# Identification of a specific inflammatory protein biosignature in coronary and peripheral blood associated with increased risk of future cardiovascular events

**DOI:** 10.1101/2023.04.06.23288168

**Authors:** Diane Proudfoot, Bruna Gigante, Nick E.J. West, Stephen P. Hoole, Rona J. Strawbridge, Elena Tremoli, Damiano Baldassarre, Simon Williams

## Abstract

**Background and rationale:** As an adjunct to coronary intervention, the Liquid Biopsy System (LBS, PlaqueTec, UK) enables accurate intracoronary blood sampling at discrete sites simultaneously. We investigated variation between local coronary and remote (peripheral) blood levels of a panel of atherosclerosis-associated proteins and examined how this might relate to cardiovascular risk assessment.

**Methods and Results:** In a previous proof-of-concept trial, coronary blood samples were collected using the LBS in 28 patients. For 12 of these patients, sampling was conducted across the uninstrumented lesion, prior to percutaneous coronary intervention (PCI). Peripheral blood samples were also collected, at baseline and after PCI. Protein levels in coronary and peripheral plasma samples were analysed by proximity extension assay (PEA, Olink).

Before PCI, in 10 out of 12 patients, coronary levels of hepatocyte growth factor (HGF), pappalysin-1 (PAPPA) and spondin-1 (SPON1) were elevated compared with peripheral levels, in some cases >10-fold. Following PCI, involving iatrogenic plaque rupture prior to stenting, peripheral levels of these proteins were elevated to a similar degree as coronary levels. In 2 patients, peripheral elevations of HGF, PAPPA and SPON1 (all >90^th^ centile) were observed at baseline, prior to PCI. The protein pattern that was identified, consisting of high levels of a combination of HGF, PAPPA and SPON1 was absent in healthy control peripheral blood, but when investigated in baseline peripheral blood samples from reference cardiovascular and COVID-19 patient cohorts, was associated with the occurrence of major adverse cardiovascular events (MACE) and mortality.

**Conclusions:** From investigation of coronary and peripheral blood samples, we identified a novel inflammatory protein signature, which when present in peripheral blood appears to portend worse outcomes. Measurement of these proteins could therefore aid identification of individuals at high risk of cardiovascular events or death.

**Translational Perspective:** Through sampling of local coronary blood, we discovered a novel protein biosignature consisting of a combination of elevated levels of HGF, PAPPA and SPON1. When this biosignature was assessed in peripheral samples from reference cardiovascular and COVID-19 cohorts, it associated with the occurrence of MACE and mortality. The biosignature protein levels correlated with markers of mast cell and neutrophil activity but not with CRP, possibly indicating a specific inflammatory status. Early detection of this protein signal has potential clinical utility to identify specific patients at increased risk of poor outcomes.

**Graphical Abstract:** 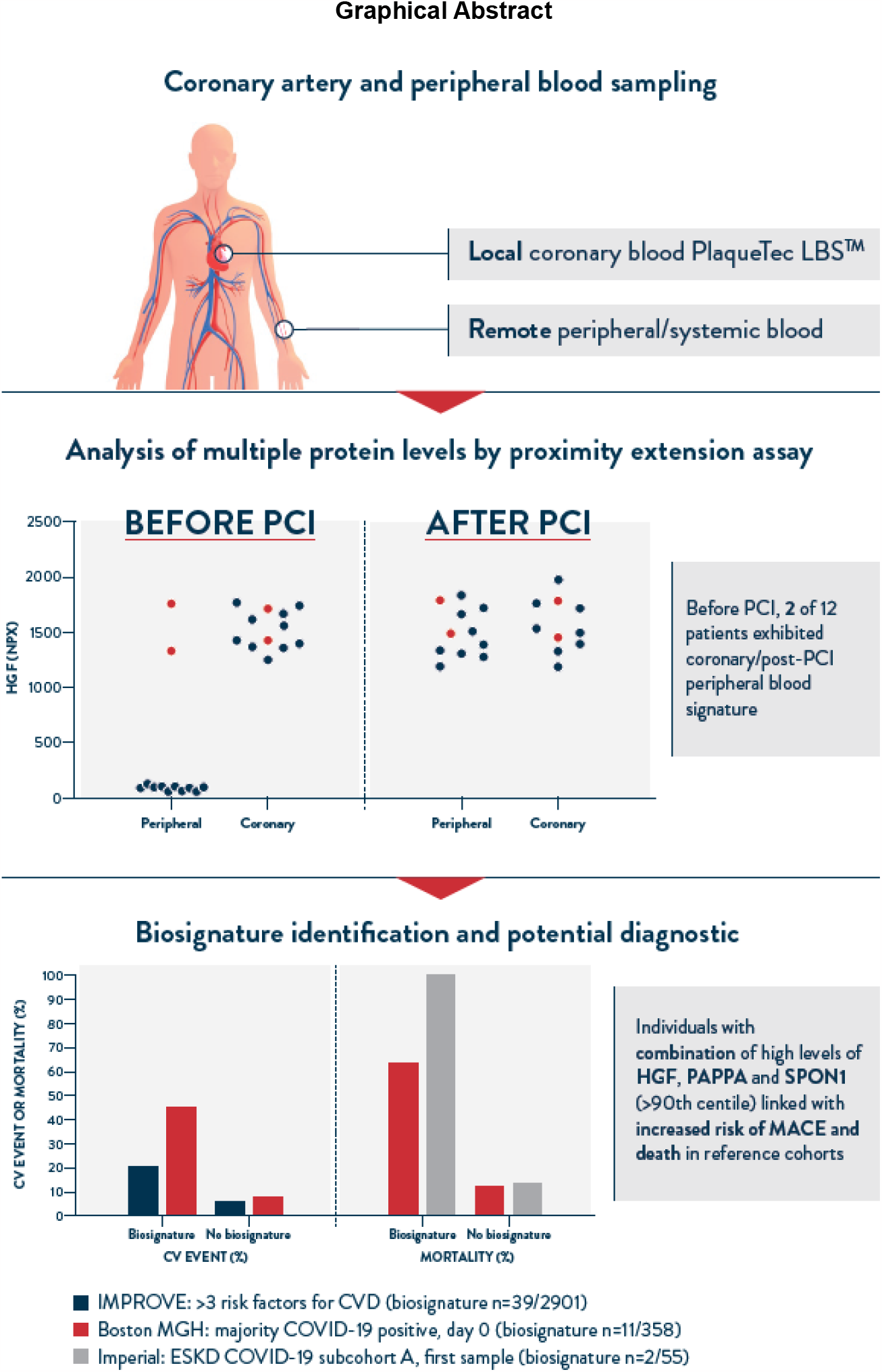

## Introduction

Despite current primary and secondary prevention strategies, coronary artery disease remains the principal cause of death and disability worldwide, indicating much is still to be understood about the mechanisms underlying residual cardiovascular risk (1, 2). Genetic insights have suggested numerous proteins, lipids, and metabolites as identifiers of increased risk (3-6) and several studies have identified circulating biomolecules associated with risk of major adverse coronary events (MACE). Principally, these studies analysed systemic venous biomolecule levels and correlated with measures of disease severity (7) or retrospective analyses of outcomes (8, 9).

Inflammation is known to contribute to atherosclerotic disease progression, destabilising plaques and promoting thrombosis; however, the specific causal components of these inflammatory pathways and their links to other factors such as shear stress, wall strain and flow disturbance are only beginning to be understood (10). Assessment of systemic inflammation by measuring C-reactive protein (CRP) levels (an acute phase reactant) has been linked with increased risk of myocardial infarction (11), and may be useful in stratifying patients most likely to benefit from targeted anti-inflammatory treatments (12). Encouragingly, large-scale randomised trials of both colchicine (13) and canakinumab (14), an inhibitor of the inflammatory mediator IL-1β, have now demonstrated a reduction in cardiovascular events vs placebo in patients with a history of myocardial infarction, demonstrating that modulating inflammation can have manifest effects on risk of clinically important outcomes in atherosclerotic vascular disease.

Systemic venous blood sampling reflects biomolecules derived from multiple organ systems and vascular beds, making the identification of specific plaque-released factors problematic. Methods used to address this have included *ex vivo* sampling of atherosclerotic plaques (15, 16) and coronary thrombi (17, 18) or use of aspiration or guide catheters to sample blood directly from the coronary artery (19-21). Samples collected from the coronary sinus, a venous reservoir receiving drainage from the majority of the coronary circulation, have been compared to blood sampled from the coronary ostium to identify transmyocardial gradients of certain proteins, potentially reflecting coronary disease status more accurately than analysis solely of systemic proteins sampled remotely (22).

The present study describes a novel protein pattern discovered in new analysis of coronary and peripheral protein data from our previous study, where intracoronary samples were collected using the PlaqueTec Liquid Biopsy System (LBS) (23). We discovered not only specific elevated signals in the coronary samples but also, in a minority of individuals, in peripheral samples from a baseline blood draw. We then searched other published cohorts for patients exhibiting this peripheral biosignature to assess prevalence and possible links to clinical outcomes.

## Materials and Methods

### Coronary artery blood sample collection

As part of a proof of concept clinical trial of the LBS device (Clinical Study 1 – CS1, ClinicalTrials.gov: NCT02119767), intracoronary samples were obtained via sampling ports positioned either side of an identified plaque in 28 patients (23). As described previously, all participants provided informed written consent; ethical approval for the study was granted by NRES London, Chelsea 13/LO/0954 (23). This analysis focused on the 12 patients from the CS1 study that underwent blood sampling prior to PCI. Peripheral blood samples were taken at baseline, prior to heparin administration, and also after PCI. LBS-derived and peripheral samples were mixed with EDTA, and plasma isolated by centrifugation to generate platelet-poor plasma and stored at -80 °C until analysis, as described previously (23).

Plasma samples were analysed by Proximity Extension Assay (PEA; Olink, Uppsala, Sweden), initially using the CVD1 panel (discontinued) as described previously (23). Separately, plasma samples were reanalysed using the Cardiovascular 3 (CVD3) and Inflammation panels, each containing 92 proteins. The full list of proteins included in the analysis are detailed in the Supplementary materials (File S1). Results are presented as normalised protein expression (NPX) units, and log2 NPX values were converted into linear scale (using 2^NPX^=linear NPX). Assay characteristics with detection limits, validations and explanations of NPX units are available from the manufacturer (http://www.olink.com). Age- and sex-matched healthy control systemic blood samples (BioIVT, Westbury, NY, USA) were analysed on the CVD3 and Inflammation panels. CRP levels were analysed by high sensitivity CRP (hsCRP) ELISA (Invitron, cat # IV3-105E; Eurofins, Abingdon, UK) and results provided in mg/L.

### Reference populations

To investigate prevalence and importance of the biosignature identified in CS1, other published studies where the same proteins have been measured by PEA (Olink) were investigated. A short summary of each cohort investigated is shown in Table 3, and details are provided below.

### PACIFIC cohort

Plasma protein data from a study by Bom et al (7) was accessed from the journal website. This study included 196 participants from the PACIFIC cohort with suspected cardiovascular disease (CVD) who underwent coronary computed tomography angiography (CCTA). Proteins were measured by PEA (Olink) using the CVD2, CVD3 and Inflammation panels.

### Healthy control cohort (Olink)

Data on normal ranges of plasma proteins measured by PEA using Olink Explore 3072 for 300 healthy individuals was accessed at the manufacturer’s website (https://insight.olink.com/data-stories/normal-ranges).

### IMPROVE cohort

The IMPROVE cohort recruited 3711 participants with at least 3 established CVD risk factors (men, post-menopausal women, dyslipidaemia, hypertension, diabetes, smoking and family history of CVD) (24, 25). Proteomic analysis of plasma was previously measured in IMPROVE using the CVD1 panel (Olink Proteomics, Uppsala, Sweden) (26), and data on levels of hepatocyte growth factor (HGF), Pappalysin-1 (PAPPA) and Spondin-1 (SPON1) were extracted. Carotid-intima media thickness (c-IMT) was measured as previously described(25) and carotid plaques, defined as c-IMT≥1.5mm; all subjects were followed to 3 years and major adverse events recorded. Because of changing rules at some institutions 491 individuals from one center in Finland have been excluded from the analysis. After quality control, 2901 participants had baseline protein and carotid measurements included in the analysis and followed up after 3 years. 171 cardiovascular events were recorded at 3-year follow-up, defined as myocardial infarction, ischemic stroke, peripheral artery disease or revascularization procedures.

### Respiratory disease/COVID-19 cohort

384 patients enrolled in the emergency department during the COVID-19 peak between March and April 2020 from Boston MGH had plasma samples analysed on the Olink Explore 1536 protein panel (27). Protein data and patient outcomes for this study were accessed via the Olink website (Olink.com).

### ESKD/COVID-19 cohorts

An Imperial College study of end-stage kidney disease (ESKD) subcohort A where 55 patients tested positive and 51 tested negative for COVID-19, and from subcohort B where 46 patients tested positive and 11 tested negative for COVID-19 (28) had plasma or serum samples analysed using PEA (Olink) assays on Inflammation, Immune Response, Cardiometabolic, Cardiovascular 2, and Cardiovascular 3 panels. Protein data and patient outcomes for this study were accessed via the journal website and personal communication with the authors.

### Statistical analysis

In the CS1 cohort, to test for statistical significance between protein levels in plasma sampled at different locations (peripheral v coronary, and pre- and post-PCI), a one-way ANOVA mixed-effects model, followed by a pairwise Holm-Sidak’s multiple comparison’s test was used (with an alpha of 0.05). Pairwise correlation coefficients between proteins were calculated by Spearman (rho coefficient) performed in GraphPad (version 9.0). In the respiratory disease COVID-19 cohort, survival analysis, Kaplan-Meier curves and statistics were created in GraphPad (version 9.0).

For the IMPROVE cohort, continuous variables are expressed as median and interquartile range, categorical variables as number and percentages. HGF, PAPPA and SPON1 data were standardized using the z-score. The correlation coefficient between the three biomarkers (pairwise) and of each one of the three biomarkers with CRP (mg/L) was estimated by Spearman’s (rho coefficient). Participants were categorized as two groups or “scores”: “0” if HGF, PAPPA and SPON1 were all <90th centile and “1” if HGF, PAPPA and SPON1 were all ≥90th centile. Kruskall Wallis and chi square tests were used to analyze differences across groups. A linear regression analysis was performed using the log transformed measures of c-IMT as dependent variable and the single biomarker or the biomarker score as independent variables. A Cox regression model was used for event analysis. Data were adjusted by latitude (main determinant of c-IMT in the IMPROVE cohort), age, gender and analytical batch. Estimates were expressed as β coefficient and standard error (SE). All analyses were performed in STATA v14.

## Results

### Identification of protein biosignature in coronary and peripheral blood samples

12 patients (45-74 years, all male, Table 1) underwent blood sampling prior to any balloon dilatation of an obstructive coronary plaque as part of a proof-of-concept clinical trial of the LBS)(23); 11 of the 12 subsequently underwent PCI.

**Table 1.**
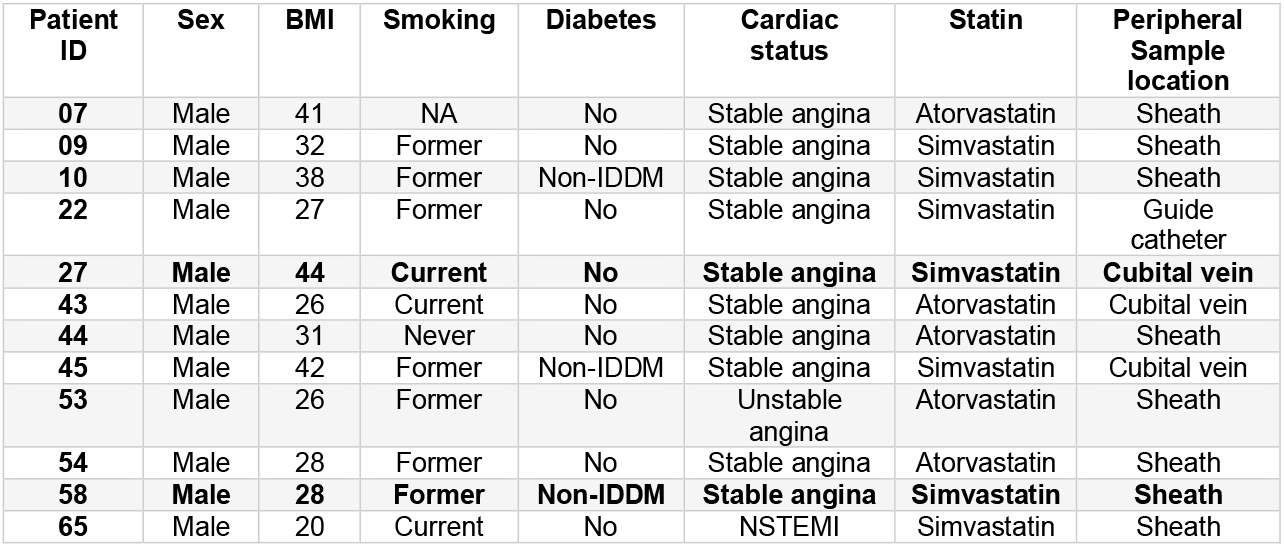
Demographic characteristics for the PlaqueTec CS1 trial. All patients with blood samples collected before PCI are listed. Locations of the blood sample taken peripherally, prior to heparin infusion. Note details for the 2 patients with the biosignature at baseline, 27 and 58, are displayed in bold. Abbreviations: insulin-dependent diabetes, IDDM; Non-ST-elevation myocardial infarction, NSTEMI.

Using the CVD1 protein panel, levels of HGF and PAPPA were identified at much higher levels (>10-fold) in the coronary samples compared with peripheral samples for 10/12 individuals (Figure 1A, Supplement Figure S1). SPON1 levels were also higher in coronary samples compared with periphery in the same 10 patients but to a lesser extent than HGF and PAPPA. The remaining 2 patients were found to have elevated levels of HGF, PAPPA and SPON1 in peripheral *as well as* coronary samples (Figure 1B, C and D), despite no differences in demographics or sample location that may have explained these differences (Table 1). Other coronary/peripheral protein peaks (Figure 1A) were not explored further as they did not have the distinct patient pattern observed with absolute levels of HGF, PAPPA and SPON1.

**Figure 1.**
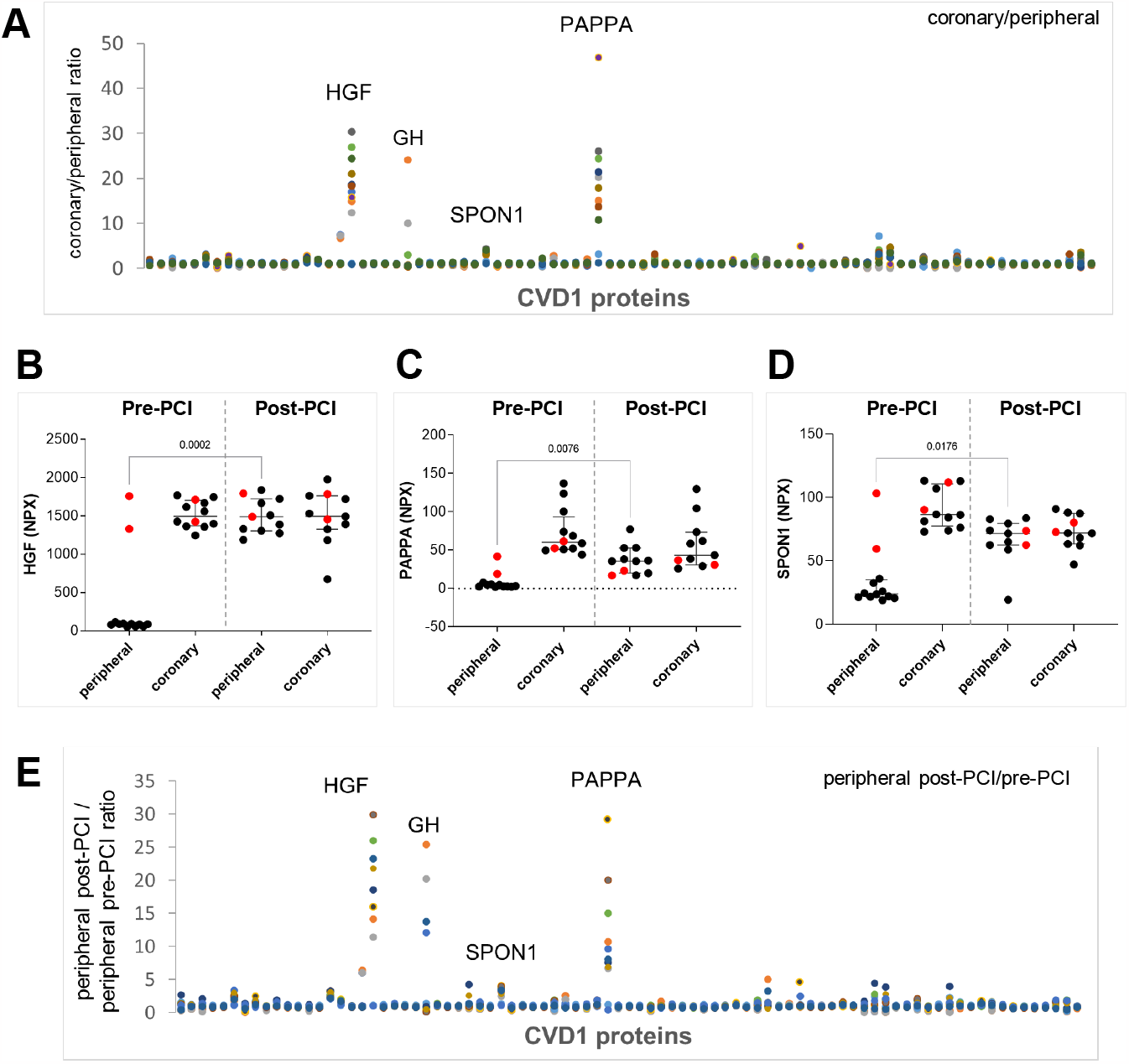
Comparison of coronary and peripheral protein levels – CVD1 protein panel. A. Protein levels in plasma samples from 12 patients from the CS1 cohort, expressed as ratio of coronary/peripheral levels (CVD1 panel proteins on x-axis). Each datapoint represents an individual patient, with each patient displayed by a different colour. Data from proximal coronary plasma samples is shown (see Supplement Figure 1 for distal coronary data). Note 3 proteins indicated have high coronary/peripheral protein ratios for 10 of the 12 patients. B. HGF absolute protein levels (NPX units) in coronary and peripheral plasma, pre- (n=12) and post-PCI procedure (n=11). Each datapoint represents an individual patient. Note that 2 patients had much higher levels of HGF (shown in red) in peripheral samples prior to PCI compared with the other 10 patients. C. As for B with absolute levels of PAPPA (NPX units). Note that 2 patients had much higher levels of PAPPA (shown in red) in peripheral samples prior to PCI compared with the other 10 patients. D. As for C with absolute levels of SPON1 (NPX units). Note that 2 patients had much higher levels of SPON1 (shown in red) in peripheral samples prior to PCI compared with the other 10 patients. E. Post-PCI/pre-PCI ratio of systemic samples (CVD1 panel). Growth Hormone (GH). Note similar patterns to A, indicating that for 10 of the 12 patients, conditions of the PCI procedure raised the peripheral levels of these specific proteins. For 2 patients, these proteins were already at high levels peripherally at baseline, before the procedure.

In post-PCI samples, the majority of patients had higher levels of HGF, PAPPA and SPON1 peripherally (Figure 1E) compared with pre-PCI samples, suggesting that the conditions of the PCI, including iatrogenic plaque rupture and resultant response to injury - inflammation and release of plaque constituents – caused a systemic elevation of these proteins.

### Confirmation of protein biosignature in coronary and peripheral blood samples

Initially considered outliers, samples from the 2 patients with high peripheral levels of HGF, PAPPA and SPON1 prior to PCI were re-tested using the ‘Inflammation’ and ‘CVD3’ panels (Olink), which include HGF and SPON1 from the now-discontinued CVD1 panel but do not include PAPPA. Plasma samples from the 10 individuals with low baseline but elevated post-PCI biosignature and from 12 healthy individuals (age- and sex-matched) as a control series were also included in these assays for comparison.

Repeat testing of both HGF and SPON1 demonstrated consistent results (Figure 2A and B). The 10 patients with low HGF and SPON1 in the peripheral samples prior to PCI had similar levels to healthy controls, indicating that the elevated levels seen in 2 patients were well above ‘normal’. Of note, some differences between absolute NPX values measured on CVD1 and CVD3/Inflammation panels were seen, most likely attributed to the different dilutions and dynamic ranges of the newer panel assays. Analysis of coronary/peripheral protein ratios using the two new panels revealed 5 further proteins with distinctly higher levels in the coronary plasma compared with peripheral plasma; C-X-C motif chemokine 9 (CXCL9), C-C motif chemokine ligand 28 (CCL28), TNF-related weak inducer of apoptosis (TNFSF12 or TWEAK) (Figure 2C), tissue factor pathway inhibitor (TFPI) and azurocidin-1 (AZU1) (Figure 2D).

**Figure 2.**
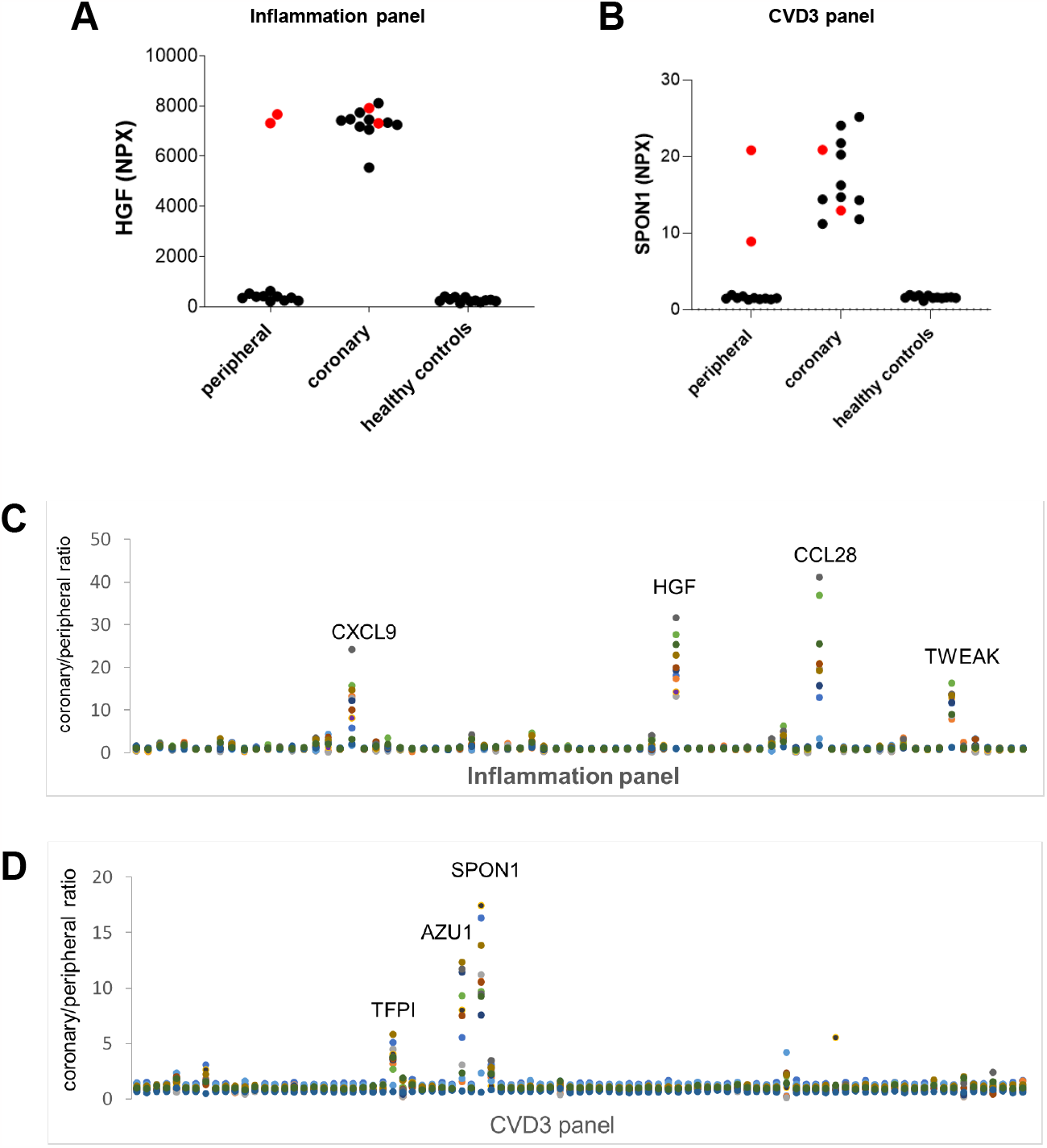
Comparison of coronary and peripheral protein levels repeat analysis of CS1 plasma in CVD3 and Inflammation protein panels. A. Repeatability measurements of CS1 coronary and peripheral samples for HGF in Inflammation panel and comparison with healthy controls (age- and sex-matched for the CS1 cohort). Note the 2 patients with much higher peripheral levels of HGF and SPON1 are shown in red. B. As for A with but with SPON1 using the CVD3 panel. (Note that patterns for HGF and SPON1 measured on the CVD1 panels in Fig 1B and D were similar, but NPX values differed between the 2 panels due to different dilution factors and dynamic ranges of the assays. C. Coronary:peripheral protein ratios, Inflammation panel. Comparing with Figure 1A, a similar HGF pattern was observed. Further proteins with similar patterns were identified: CXCL9, CCL28 and TWEAK. D. Coronary:peripheral protein level ratios, CVD3 panel. Comparing with Figure 1A, a similar but more distinct SPON1 pattern was observed. Further proteins with similar patterns were identified: TFPI and AZU1.

When comparing absolute coronary and peripheral protein values a distinct pattern was seen for HGF, which to varying extent was mirrored by the 7 other proteins. The 2 patients with the biosignature had high levels of AZU1 and CXCL9, however, one other patient in each case had high peripheral levels (Figure 3A). Some healthy control participants had relatively high levels of AZU1 and CXCL9. The peripheral biosignature pattern appeared to be most consistent for 6 proteins: HGF, PAPPA, SPON1, CCL28, TFPI and TWEAK.

**Figure 3.**
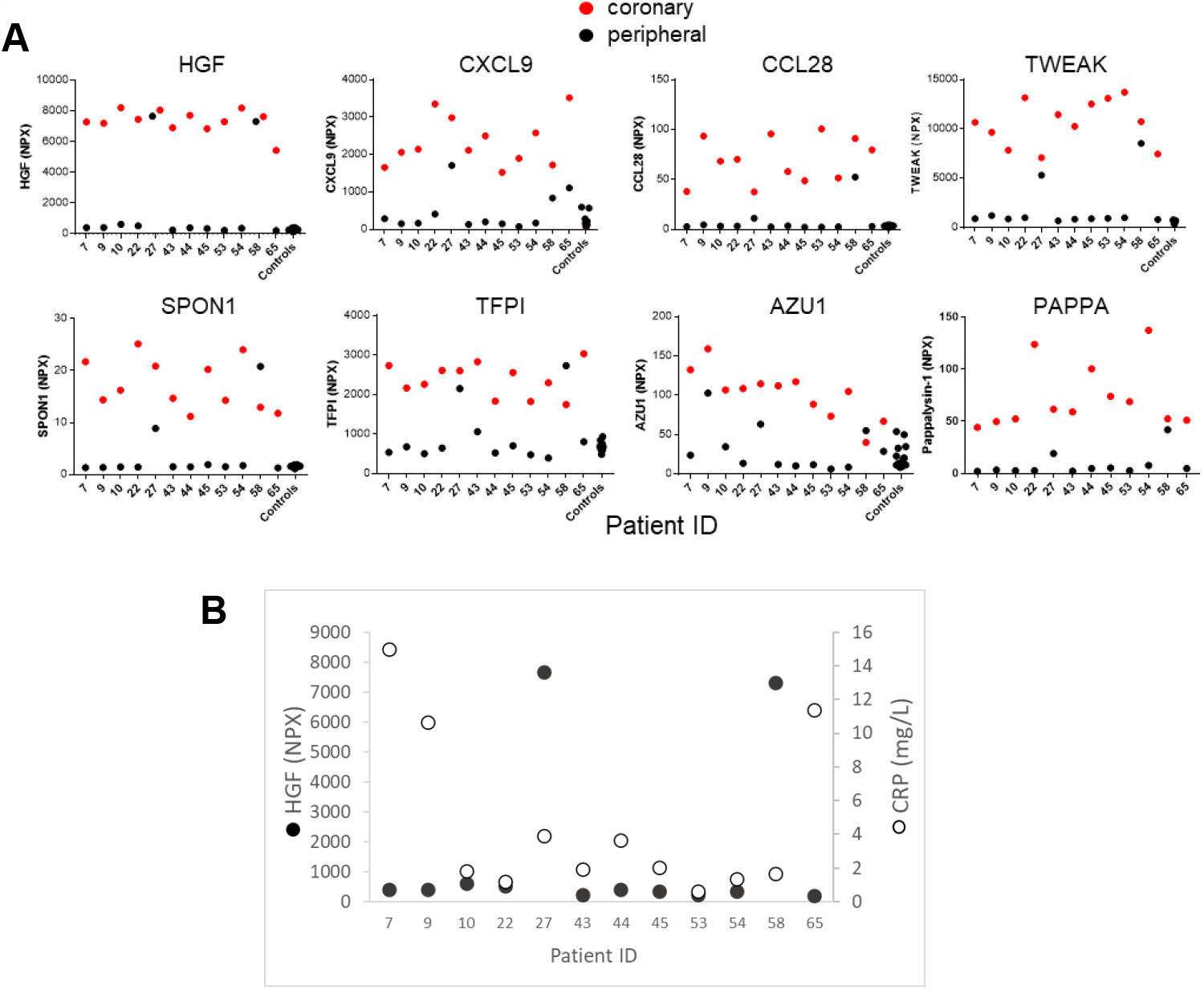
CS1 patient and healthy control peripheral proteins levels. A. Individual patient absolute levels (NPX) for selected proteins, coronary (red) and peripheral (black), and comparing with 12 healthy, age- and sex-matched control plasma samples, data from the CVD3 and Inflammation panels. These raw data plots revealed that levels of these proteins in coronary arteries of patients are clearly higher than healthy, age- and sex-matched control peripheral levels (n=12). This effect was less pronounced for AZU1 and CXCL9. PAPPA control levels were not determined as PAPPA is not included on the CVD3 and Inflammation panels. Thus 2 patients had a clear systemic pattern of elevated HGF, SPON1, TFPI, CCL28, TWEAK and PAPPA. B. No correlation of hsCRP with HGF levels Spearman R = -0.01 (P = 0.99).

### Analysis of C-Reactive Protein (CRP) levels

To investigate whether the peripheral biosignature pattern seen in 2 patients could be explained by acute-phase systemic inflammation, CRP levels were measured using a high-sensitivity assay (hsCRP). Levels were uniformly low (<5mg/L), and CRP did not correlate with HGF levels (Figure 3B), suggesting systemic inflammation was not a factor.

### Investigating the peripheral biosignature in healthy individuals

To further investigate the biosignature in healthy cohorts, data available from Olink on variability (IQR) of 2943 proteins in a healthy cohort of 300 individuals, the 6 biosignature proteins had relatively low variability, compared with in the PACIFIC and CS1 cohorts (Supplement Figure S2). If a minority of participants had unusually high levels of the 6 proteins, this would raise IQR values relative to the other proteins measured. Observing relatively low IQRs for the 6 proteins in this cohort suggests it was unlikely that any of the healthy participants had the biosignature.

### Investigating the peripheral biosignature pattern in patients with suspected coronary artery disease: PACIFIC cohort

A study by Bom et al (7) represents a 196-patient subset of the PACIFIC cohort with suspected coronary artery disease. Two of the 196 patients displayed clearly raised levels of all six biosignature proteins (Figure 4). To allow comparisons across different cohorts, a biosignature definition of >90^th^ centile for HGF and also PAPPA and SPON1 was used, which identified three participants with the biosignature, all with low CRP levels (hsCRP<2.5mg/L). None of the demographic variables provided clues to differentiate the biosignature-displaying subjects from the rest of the cohort.

**Figure 4.**
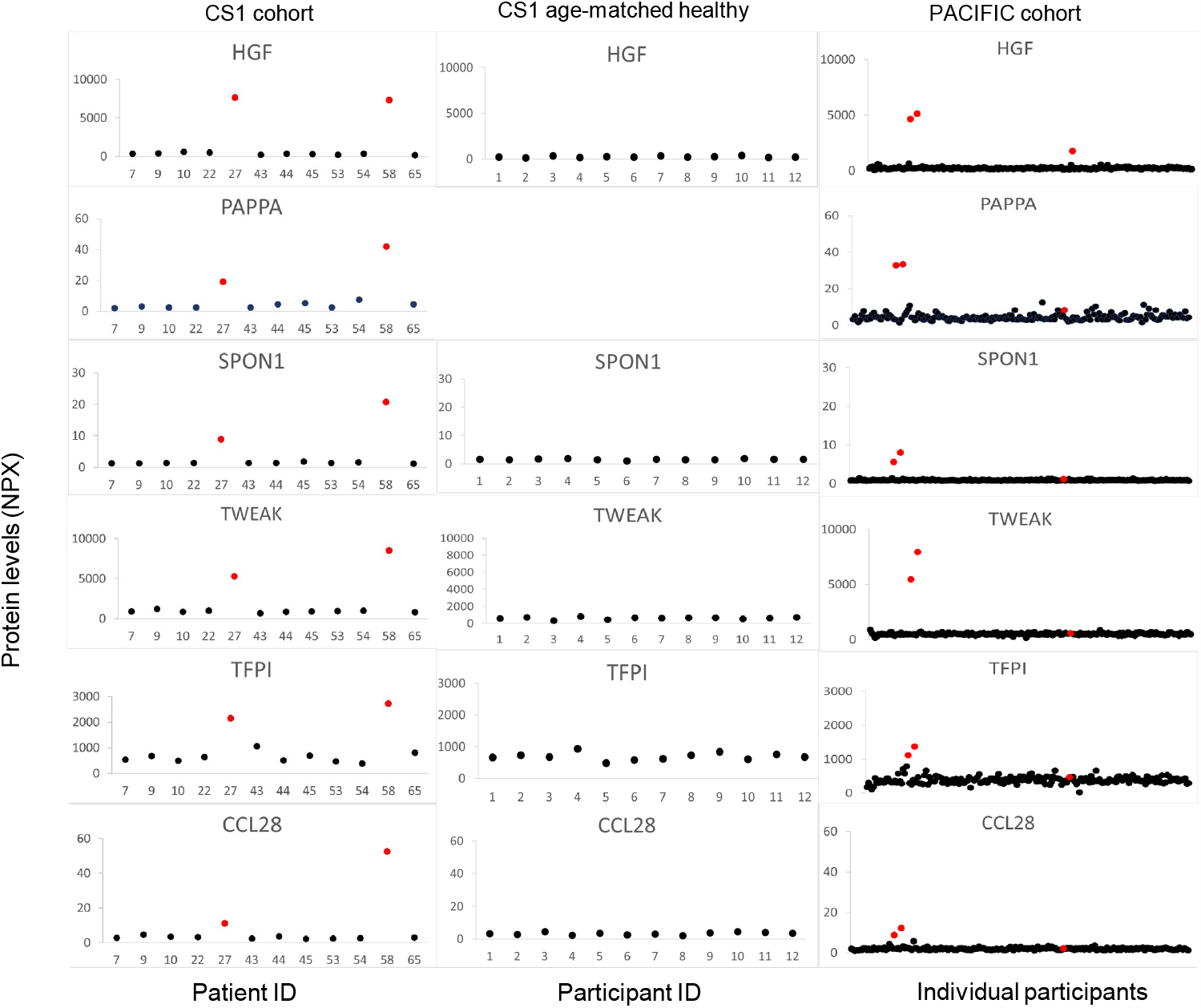
Comparison of CS1 with PACIFIC cohort. Absolute levels for peripheral proteins are displayed for CS1 cohort (left) and data was downloaded from Bom et al^7^, PACIFIC cohort (right). Age-matched control levels can be compared with CS1 cohort as they were analysed together (PAPPA was not included). NPX levels between CS1 and PACIFIC cohorts cannot be compared directly, but patterns can be identified, displaying similarities in the expression levels of the 6 proteins. 2 participants in the PACIFIC cohort displayed distinctly higher levels of the 6 proteins compared with the majority of participants (indicated in red). Using a definition of >90^th^ centile for HGF and also PAPPA and SPON1 identified 1 additional participant (indicated in red).

### Investigating the peripheral biosignature pattern in patients with multiple CVD risk factors: IMPROVE cohort

In the IMPROVE population, three of the biosignature proteins were measured: HGF, PAPPA and SPON1. 39 subjects were identified with the biosignature defined as >90th centile levels for HGF, and also PAPPA and SPON1. This was a small minority (1.3%) of the cohort that comprised 26 men and 13 women (Table 2A). The signature proteins associated with a thicker c-IMTmax (P=0.033) (Table 2B). The biosignature proteins displayed a positive and significant correlation with each other (Table 2C). No correlation was observed between biosignature proteins and CRP levels, except for HGF. However, the biosignature group had higher CRP levels compared with the remaining cohort (Table 2A). During the 3-year follow-up, 20.5% (8 of 39) of the patients with the biosignature suffered a cardiovascular event, compared with 5.7% (163 of 2862) in the remaining cohort. Using a Cox regression model, participants with the biosignature had a significantly increased risk of future cardiovascular events; HR 3.16 (95%CI:1.55-6.45), p=0.002. Additionally, when applying a 90^th^ percentile definition for identification of participants with the highest levels of individual proteins and analysing event rates for HGF (15 of 171, 8.8%), PAPPA (17 of 171, 9.9%) or SPON1 alone (20 of 171, 11.7%), suggested that the biosignature combination of proteins performs better at predicting MACE compared with each protein analysed independently. Furthermore, for participants with CRP above the 90^th^ percentile, 28/171 had an event (16.4%) which suggests that the biosignature is also better than CRP at detecting the occurrence of MACE.

**Table 2.**
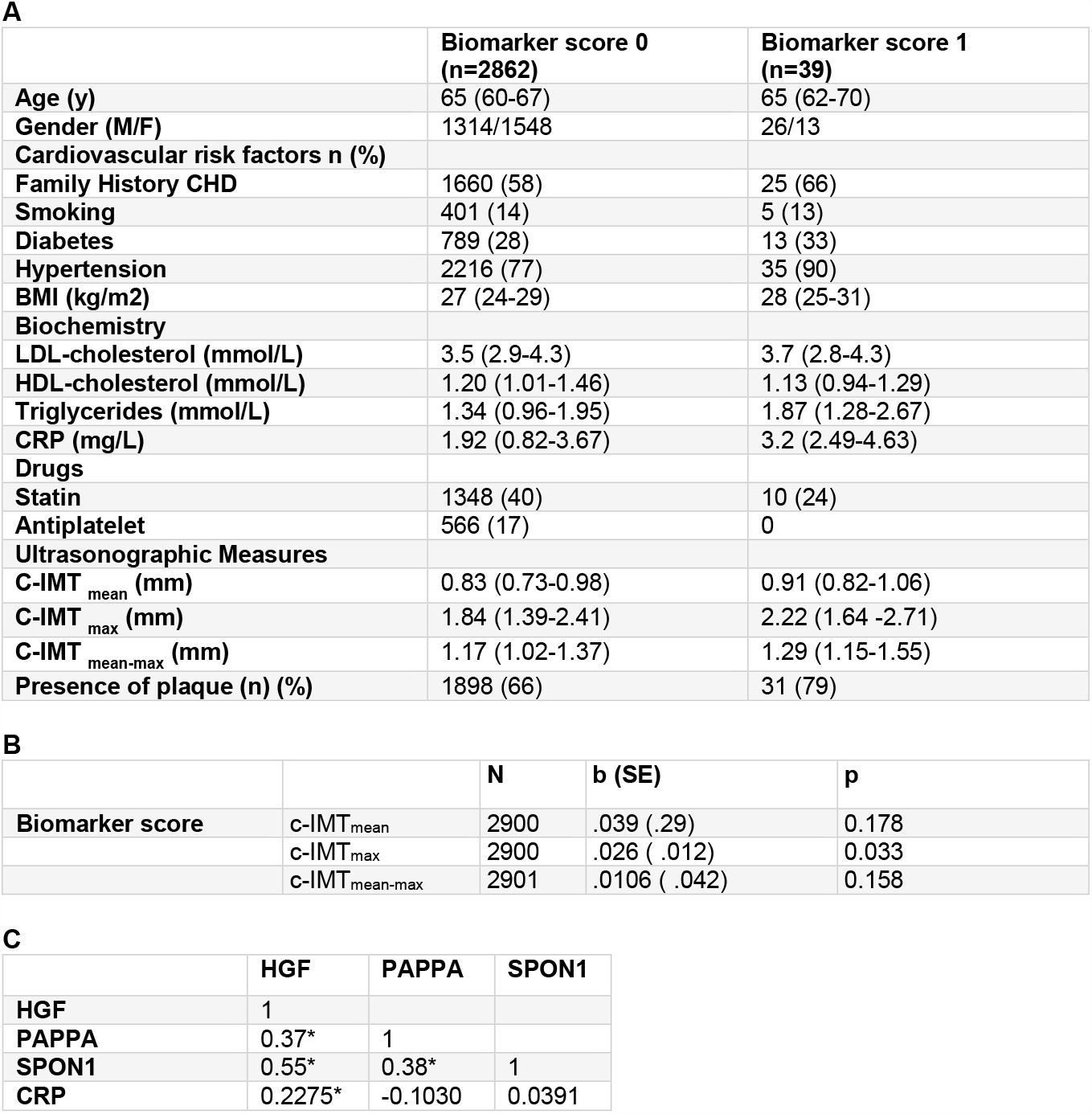
IMPROVE cohort and biosignature. A. Demographic characteristics for IMPROVE according to the plasma levels for HGF, and also PAPPA and SPON1 included in the biomarker score categorized as below (0) and over (1) the 90^th^ percentile for all three proteins. Continuous variables are reported as median (interquartile range) and categorial variables as number (percentages); CHD: coronary heart disease; Presence of plaque: number of study participants where a plaque has been recorded. B. Association of the biomarker score 1 (> 90^th^ percentile) with measures of c-IMT mean or maximum (max) after adjustment by age, gender and latitude by linear regression. (Note that latitude is highly correlated with c-IMT and plaque). C. Pairwise correlation between the biomarkers in analysis expressed by Spearman rho coefficient, *\* p<0*.*0001*

|  | **Biomarker score 0**<br>**(n=2862)** | **Biomarker score 1**<br>**(n=39)** |
| --- | --- | --- |
| **Age (y)** | 65 (60-67) | 65 (62-70) |
| **Gender (M/F)** | 1314/1548 | 26/13 |
| **Cardiovascular risk factors n (%)** |  |  |
| **Family History CHD** | 1660 (58) | 25 (66) |
| **Smoking** | 401 (14) | 5 (13) |
| **Diabetes** | 789 (28) | 13 (33) |
| **Hypertension** | 2216 (77) | 35 (90) |
| **BMI (kg/m2)** | 27 (24-29) | 28 (25-31) |
| **Biochemistry** |  |  |
| **LDL-cholesterol (mmol/L)** | 3.5 (2.9-4.3) | 3.7 (2.8-4.3) |
| **HDL-cholesterol (mmol/L)** | 1.20 (1.01-1.46) | 1.13 (0.94-1.29) |
| **Triglycerides (mmol/L)** | 1.34 (0.96-1.95) | 1.87 (1.28-2.67) |
| **CRP (mg/L)** | 1.92 (0.82-3.67) | 3.2 (2.49-4.63) |
| **Drugs** |  |  |
| **Statin** | 1348 (40) | 10 (24) |
| **Antiplatelet** | 566 (17) | 0 |
| **Ultrasonographic Measures** |  |  |
| **C-IMT<sub>mean</sub> (mm)** | 0.83 (0.73-0.98) | 0.91 (0.82-1.06) |
| **C-IMT<sub>max</sub> (mm)** | 1.84 (1.39-2.41) | 2.22 (1.64 -2.71) |
| **C-IMT<sub>mean-max</sub> (mm)** | 1.17 (1.02-1.37) | 1.29 (1.15-1.55) |
| **Presence of plaque (n) (%)** | 1898 (66) | 31 (79) |

B
|  |  | **N** | **b (SE)** | **p** |
| --- | --- | --- | --- | --- |
| **Biomarker score** | c-IMT<sub>mean</sub> | 2900 | .039 (.29) | 0.178 |
|  | c-IMT<sub>max</sub> | 2900 | .026 ( .012) | 0.033 |
|  | c-IMT<sub>mean-max</sub> | 2901 | .0106 ( .042) | 0.158 |

**Table 2. IMPROVE cohort and biosignature**
|  | **HGF** | **PAPPA** | **SPON1** |
| --- | --- | --- | --- |
| **HGF** | 1 |  |  |
| **PAPPA** | 0.37* | 1 |  |
| **SPON1** | 0.55* | 0.38* | 1 |
| **CRP** | 0.2275* | -0.1030 | 0.0391 |

**Table 3.**
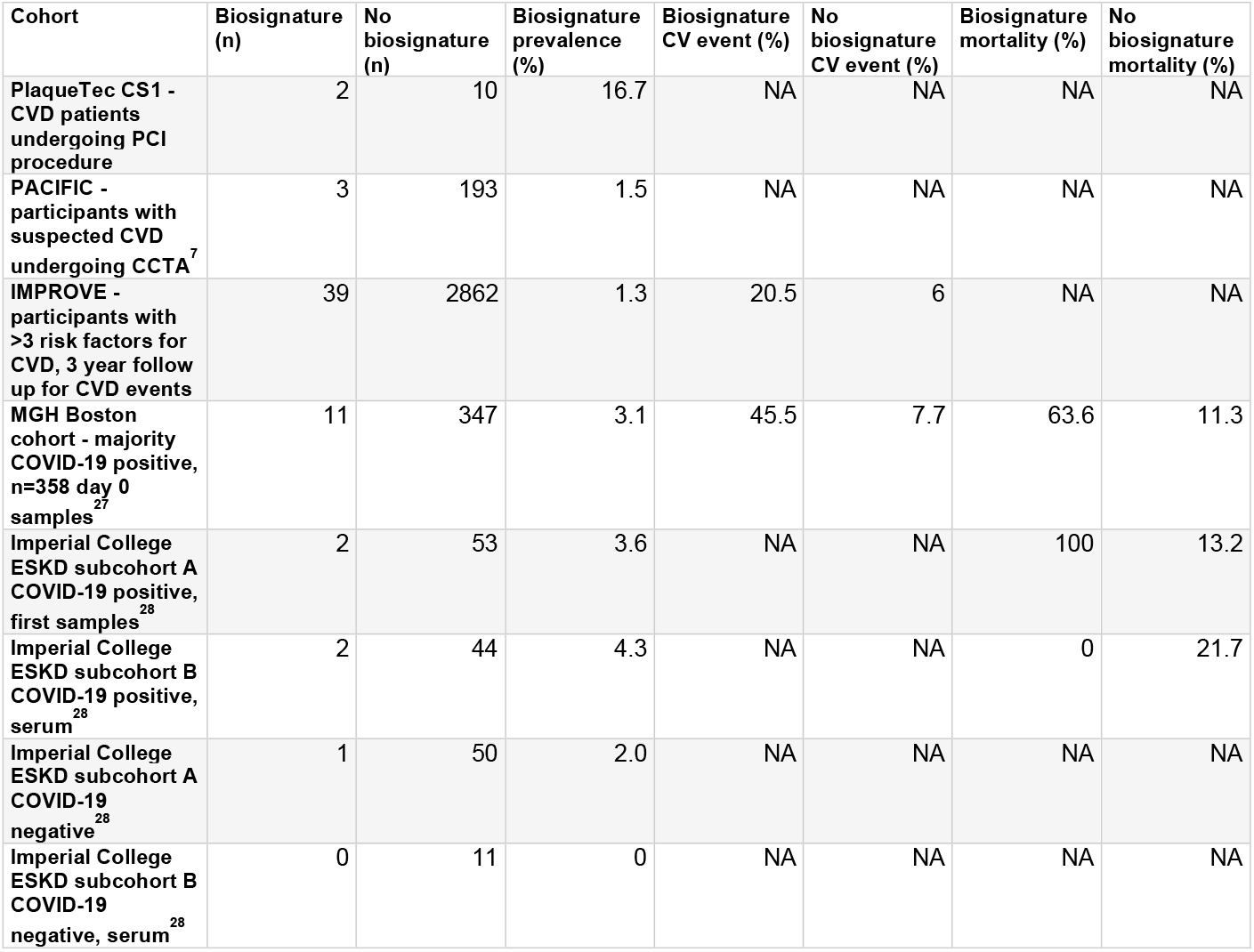
Summary of biosignature prevalence and outcomes in different cohorts. The table displays comparisons of biosignature prevalence and outcomes between cardiovascular disease cohorts, a respiratory disease/COVID-19 cohort and an ESKD/COVID-19 cohort. To identify participants with the biosignature a definition of >90th centile for HGF, and also >90th centile for PAPPA and SPON1 levels was used. Individuals identified with the biosignature were in the minority, but a had higher incidence of cardiovascular events in IMPROVE and the respiratory disease/COVID-19 cohort, and a higher mortality in the respiratory disease/COVID-19 cohort and the ESKD/COVID-19 subcohort A compared with individuals without the biosignature. Abbreviations: CV, cardiovascular; CVD, cardiovascular disease; CCTA, coronary computed tomography angiography; ESKD, end-stage kidney disease; NA, not available.

| Cohort | Biosignature (n) | No biosignature (n) | Biosignature prevalence (%) | Biosignature CV event (%) | No biosignature CV event (%) | Biosignature mortality (%) | No biosignature mortality (%) |
| --- | --- | --- | --- | --- | --- | --- | --- |
| PlaqueTec CS1 - CVD patients undergoing PCI procedure | 2 | 10 | 16.7 | NA | NA | NA | NA |
| PACIFIC - participants with suspected CVD undergoing CCTA<sup>7</sup> | 3 | 193 | 1.5 | NA | NA | NA | NA |
| IMPROVE - participants with >3 risk factors for CVD, 3 year follow up for CVD events | 39 | 2862 | 1.3 | 20.5 | 6 | NA | NA |
| MGH Boston cohort - majority COVID-19 positive, n=358 day 0 samples<sup>27</sup> | 11 | 347 | 3.1 | 45.5 | 7.7 | 63.6 | 11.3 |
| Imperial College ESKD subcohort A COVID-19 positive, first samples<sup>28</sup> | 2 | 53 | 3.6 | NA | NA | 100 | 13.2 |
| Imperial College ESKD subcohort B COVID-19 positive, serum<sup>28</sup> | 2 | 44 | 4.3 | NA | NA | 0 | 21.7 |
| Imperial College ESKD subcohort A COVID-19 negative<sup>28</sup> | 1 | 50 | 2.0 | NA | NA | NA | NA |
| Imperial College ESKD subcohort B COVID-19 negative, serum<sup>28</sup> | 0 | 11 | 0 | NA | NA | NA | NA |

### Investigating the peripheral biosignature pattern in patients with respiratory disease: COVID-19 cohort

The biosignature was also investigated in raw data available from the MGH Boston COVID-19 cohort (27) using the same biosignature definition to identify individuals with >90^th^ percentile values for HGF, and also PAPPA and SPON1. Analysis of protein levels at day 0 (first sample collected from 358 individuals) identified 11/358 patients (3.1%) with the biosignature, a slightly higher prevalence compared with IMPROVE (Supplement Figure S3 and Summary results in Table 3). For individuals identified with the biosignature, 7 of the 11 (63.6%) died within 28 days of admission to hospital, while 42 of the 373 (11.3%) remaining patients died. For the 4 surviving subjects with the biosignature, 2 were ‘intubated, ventilated and survived to 28 days’ and 2 were ‘hospitalized, supplementary O2 required’. Survival analysis indicated a significant statistical difference in probability of survival between those with and without the biosignature (P<0.001, Supplement Figure S4). Individuals with the biosignature had higher rates of cardiac events within the first 72 hours of presentation (5/11, 45%), compared with those without the biosignature (29/373, 8%). Participants with the biosignature also had higher rates of pre-existing kidney disease (5/11, 45%) compared with participants without the biosignature (56/373, 15%) and had similar CRP ranges to those without the biosignature (Supplement Figure S5 and Summary results in Table 3 and Table 4).

**Table 4.** Comparison of biosignature CRP levels and correlations with MPO, tryptase and syndecan-1 in different cohorts. For CRP levels, medians (range) are shown. For correlations, proteins with a positive and significant (p<0.05) Spearman pairwise correlation are shown. Results of correlation analysis are detailed in the Supplement File S1. NA, not available.

| Cohort | Biosignature CRP (mg/L) | No biosignature CRP (mg/L) | CRP higher in Biosignature group? | Biosignature proteins correlate with CRP | Biosignature proteins correlating positively with MPO | Biosignature proteins correlating positively with tryptase | Biosignature proteins correlating positively with syndecan-1 |
| --- | --- | --- | --- | --- | --- | --- | --- |
| **CVD PlaqueTec CS1** | 3.4 (2.1-4.7) | 12.8 (0.7-15) | no | no | HGF, TFPI, CCL28 | NA | NA |
| **CVD PACIFIC<sup>7</sup>** | <2.5 | <2.5 n=159<br>>2.5 n=33 | no | NA | HGF, PAPPA, SPON1, TWEAK, TFPI | NA | NA |
| **CVD IMPROVE** | 3.2 (2.49-4.63) | 1.92 (0.82-3.67) | Yes | HGF | HGF, PAPPA, SPON1 | NA | NA |
| **COVID-19<sup>27</sup>** | 0-180+<br>(Category 1-5) | 0-180+<br>(Category 1-5) | no | NA | NA | HGF, PAPPA, SPON1, TWEAK, TFPI, CCL28 | HGF, PAPPA, SPON1, TFPI, CCL28 |
| **ESKD subcohort A<sup>28</sup>** | 220.7 (163.5-278) | 79.9 (0.6-386.1) | no | HGF | HGF, PAPPA, SPON1, TWEAK, TFPI, CCL28 | HGF | NA |
| **ESKD subcohort B<sup>28</sup>** | 197.6 (34.4-360.7) | 155.6 (18.3-405.6) | no | NA | none | TWEAK, CCL28 | NA |

### Investigating the peripheral biosignature pattern in patients with multiple CVD risk factors: ESKD COVID-19 cohort

The biosignature was then investigated in an Imperial College study of 2 end-stage kidney disease (ESKD) cohorts: subcohort A and B (28). Of the patients that tested positive for COVID-19, 2/55 patients (3.6%) in subcohort A and 2/46 patients (4.3%) in subcohort B were identified with the biosignature in first samples collected (Supplement Figure S6 and Summarised in Table 3). In subcohort A, both patients with the biosignature died during the study, while in subcohort B both patients survived and were classed as having ‘severe’ or ‘moderate’ disease. In subcohort A, CRP correlated with HGF levels but not with the other biosignature proteins (28). Participants had wide ranges of CRP levels and no statistical differences were found between groups (Supplement Figure S7 and Table 4).

### Biosignature association with Tryptase, MPO, and Syndecan-1 levels

The biosignature pattern in plasma could possibly be induced via endogenous heparin release from mast cells, and/or neutrophil activity and glycocalyx damage. To test if this notion is consistent with proteins known to be linked with these processes, correlation analysis was performed between the biosignature proteins and tryptase, myeloperoxidase (MPO), and syndecan-1 levels in peripheral blood. Significant positive correlations were found between some or all of the biosignature proteins with tryptase, MPO, and syndecan-1 levels in cohorts where this data was available (Table 4, and Supplement File S1).

## Discussion

Analysis of coronary and peripheral proteins from patients with coronary artery disease undergoing PCI revealed a novel biosignature, which prompted investigation of the peripheral protein pattern in other cohorts. Using the definition of >90^th^ centile levels for HGF, and also PAPPA and SPON1, the biosignature was found in a minority of patients from a cohort of individuals with suspected coronary disease (PACIFIC cohort), and in a cohort with known CVD risk factors (IMPROVE). The biosignature was also detected in a minority of individuals in two COVID-19 cohorts but appeared absent in healthy cohorts and in healthy control plasma, which raised the question as to whether this biosignature might reflect an unstable, higher-risk cardiovascular state.

The known functions of the biosignature proteins are summarised in Table 5. All have direct or indirect roles in inflammation and endothelial function and some have been associated with risk of adverse events. For example, elevated HGF levels have been linked with worse outcomes in dialysis patients (29) and also more recently in COVID-19 cohorts, with neutrophil activation suggested as a possible source of the high HGF levels preceding critical illness (30). In a genome-wide meta-analysis of 85 proteins in over 30,000 individuals (SCALLOP study) several proteins were identified as causative in disease and included HGF (positively associated with high triglyceride levels), SPON1 (positively associated with atrial fibrillation) and PAPPA (positively associated with type 2 diabetes) (31). Interestingly, in the Imperial College study of ESKD and COVID-19 patients, SPON1 and CCL28 elevated levels were linked with increased risk of death, while increased levels of TWEAK were associated with reduced risk of death(28). TWEAK plays a role in inflammation and repair in several diseases and is thought to promote the development of atherosclerosis, but lower circulating levels have been linked to higher cardiovascular risk (32). Furthermore, 18 proteins found to be independently associated with cardiovascular death included HGF and SPON1 (9). However, the combination of specific proteins at raised levels described here has not been reported previously.

**Table 5.** Summary of functions of the biosignature proteins.

| Protein<br>(Uniprot<br>ID) | Function | Plasma levels and CVD<br>outcomes |
| --- | --- | --- |
| **HGF**<br>(P14210) | Protective role in atherosclerosis: anti-apoptotic, anti-fibrotic, pro-angiogenic, anti-inflammatory. Linked with unstable carotid atherosclerosis and elevated triglyceride levels. | High levels linked with CVD risk and mortality(9, 36)<br>Causal role in raised triglyceride levels(31)<br>Increased risk of death in COVID-19 cohort(37) |
| **PAPPA**<br>(Q13219) | Metalloprotease that cleaves IGF binding proteins, allowing IGF to bind to its receptor, which can have several effects on vascular cells including release of inflammatory cytokines. High levels linked unstable plaques and type 2 diabetes | High levels linked with CVD risk(38)<br>Causal role in Type 2 diabetes(31) |
| **SPON1**<br>(Q9HCB6) | Inhibitor of calcification in bone. Regulated by fibroblast growth factor 2 (FGF2), retinoic acid, IGF and Toll-like Receptor-4 agonists. Upregulated in osteoarthritis. Inhibitor of vascular smooth muscle cell proliferation and migration. Role in atrial fibrillation (AF). | Higher levels linked with adverse events in CHD cohort(39) and mortality(9)<br>Causal role in AF(31)<br>Increased risk of death in COVID-19 cohort(28) |
| **TWEAK**<br>(O43508) | Binds to TWEAKR inducing apoptosis, proliferation and migration of endothelial cells. Stimulates cytokine secretion and chemokine IL-8, via NFkB activation. Linked with endothelial dysfunction. | Low levels linked with higher CVD risk(40)<br>Higher levels with higher CVD risk in SLE and controls(41). Higher levels with lower death rates in COVID-19 cohort(28) |
| **TFPI**<br>(P10646) | Binds factor X and interacts with lipoproteins. Anti-thrombotic and associated with inflammation and endothelial dysfunction. | High levels linked with cardiovascular damage and myocardial infarction(42) |
| **CCL28**<br>(Q9NRJ3) | Chemokine that binds receptors CCR10 and CCR3 and exerts chemotaxis in T, B cells and eosinophils. Induced by pro-inflammatory cytokines and bacteria. | CVD not directly tested<br>Predictor of death in a COVID-19 cohort(28) |

The raised levels of biosignature proteins in the periphery may be due to local changes causing release of proteins from the endothelial glycocalyx, activated/damaged cells or activated neutrophils or mast cells, or inflammation-driven increased de novo synthesis. Of particular relevance, mast cells contain heparin (33) and mast cell activation leading to the release of endogenous heparin could increase blood levels of the biosignature proteins via release from the endothelial glycocalyx. Increased mast cell activity, as measured by serum tryptase levels, was reported as an indicator of poor CVD outcomes and tryptase levels did not correlate with CRP levels in that study, suggesting CRP is not a surrogate marker for mast cell activity (34). In the present study, individuals with the biosignature in the CVD cohorts had low-mid CRP levels (<5mg/L) which indicates a low to moderate mortality risk (35). The biosignature protein levels did not correlate with CRP levels, except in the IMPROVE and ESKD/COVID-19 cohorts where CRP correlated positively and significantly with one biosignature protein, HGF (28). However, it should be noted that all patients in the CS1 study were taking statins, which may have impacted CRP levels. In cohorts where tryptase data was available, positive correlations between biosignature proteins and tryptase were detected, suggesting there may be a link with mast cell activity. Additionally, the biosignature also appeared to be linked with neutrophil activity (MPO levels) and glycocalyx damage (Syndecan-1 levels).

To understand if an elevation in biosignature proteins is relevant to patient outcomes, we explored its presence in the IMPROVE study population of individuals at high cardiovascular risk. The biosignature group had a significant association with the risk of cardiovascular events and the biosignature protein combination appeared to be better at predicting MACE than the individual proteins or CRP. In the MGH study of patients hospitalised due to COVID-19-related symptoms (27), patients that exhibited the biosignature had higher death rates compared to patients without the biosignature. A similar observation was made in a cohort of patients with ESKD and COVID-19 (subcohort A) (28), where higher mortality rates occurred in those with the biosignature, compared with patients without the biosignature. However, in a second small ESKD COVID-19 cohort (subcohort B) (28) those with the biosignature had severe or moderate disease and survived. Some differences between cohort A and B might help to explain dissimilar results related to the biosignature, although equally the results could indicate that the biosignature may not be a reliable risk indicator for more severely ill ESKD patients.

The biosignature appears to be present in a minority of individuals, which perhaps explains why it is has not previously been described. It might have been overlooked in other studies if unusually high protein levels were considered as outliers. Furthermore, since the biosignature was discovered by comparing coronary and peripheral protein levels, the LBS sampling technology and study design may have provided a unique opportunity to make accurate coronary measurements for comparison with the periphery.

## Limitations

Although the a priori sample size for this investigation was small, the confirmation of elements of the inflammatory biosignature in much larger and well-characterised populations, particularly when correlated with increased risk of future events, is compelling. Initially, timing of sample collection and the presence of heparin had potential to explain the pre- and post-PCI disparity, however sequential comparison with other cohorts confirmed this was not the case. Although its observation in reference cohorts strengthens the validity of the biosignature, comparison of unequal groups with and without the biosignature results in wide confidence intervals. The choice of threshold of >90^th^ centile for HGF, PAPPA and SPON1 was arbitrary but aided the identification of participants with the highest NPX values for all of these proteins within a cohort and allowed comparisons across cohorts. For future applications, it would be desirable to optimise threshold selection and to measure proteins using standard concentration units (pg/ml). Nevertheless, our findings should be considered hypothesis-generating, and prospective studies on larger at-risk populations are warranted, with a focus on mechanisms underpinning the abnormally-elevated biomolecules and their role in the aetiology of cardiovascular risk.

## Conclusions

A biosignature was detected in individuals with known CVD, suspected CVD or with CVD risk factors. The biosignature associated with a higher rate of MACE, and in COVID-19 cohorts it appeared to be associated with increased risk of death. The biosignature has the potential to be developed as a non-invasive blood test indicative of a specific type of vascular inflammation and we can speculate its use as a warning signal, especially in cases where CRP levels raise no concern, and where alternative anti-inflammatory therapies may be required. An upcoming 300 patient study using the same intracoronary sampling device will enable this finding and the underlying mechanisms to be further investigated on a larger scale.

## Data Availability

For CS1 anonymised data, the majority of data presented is in raw format. Specific data requests can be considered by the corresponding author.
The IMPROVE data presented in this study are available on request from Dr. Bruna Gigante. The data are not publicly available due to ethical reasons.

## Acknowledgments

We would like to thank participants in the clinical trials detailed in this manuscript, and to the hospital staff involved in blood sample collection and processing.

We are grateful for the open access to raw data from the PACIFIC, MGH Boston and Imperial cohorts and to James Peters and Jack Gisby (Imperial College London) for additional data on subcohort B and personal communication.

This study was funded by PlaqueTec Ltd. Collaborative work on IMPROVE with BG was funded by Stiftelsen Sigurd & Elsa Goljes minne (LA2022-0133), Stiftelsen Professor Nanna Swartz fond (2022-00472) and Hjärt-Lungfonden (20210472). IMPROVE was funded by the V^th^ European Union (EU) program.

