## Supplement Figures for "Identification of a specific inflammatory protein biosignature in coronary and peripheral blood associated with increased risk of future cardiovascular events"

### Supplementary Figures

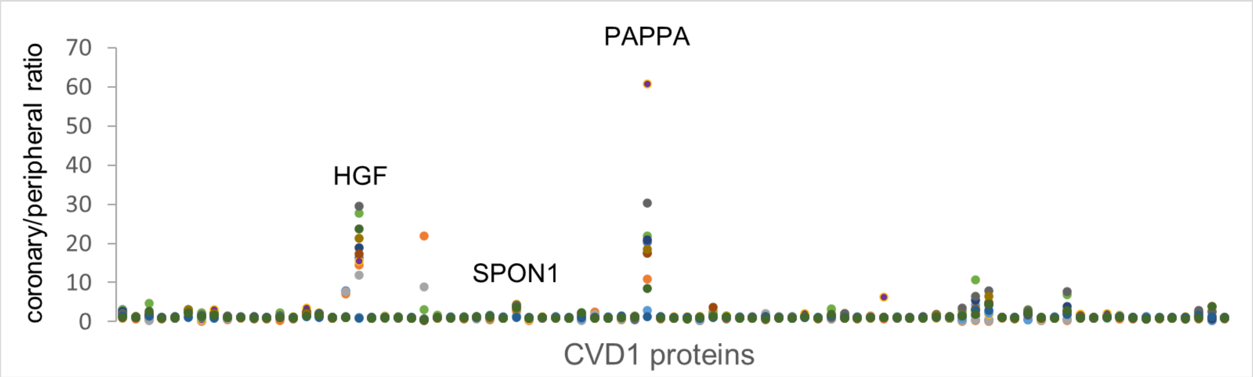


**Supplement Figure S1. Comparison of coronary and peripheral protein levels – CVD1 protein panel**
Protein levels in plasma samples from 12 patients from the CS1 cohort, expressed as ratio of coronary/peripheral levels (CVD1 panel proteins on x-axis). Data from distal coronary samples is shown. Each datapoint represents an individual patient, with each patient displayed by a different colour. Note 3 proteins indicated have high coronary/peripheral protein ratios for 10 of the 12 patients.

### Supplementary Figures

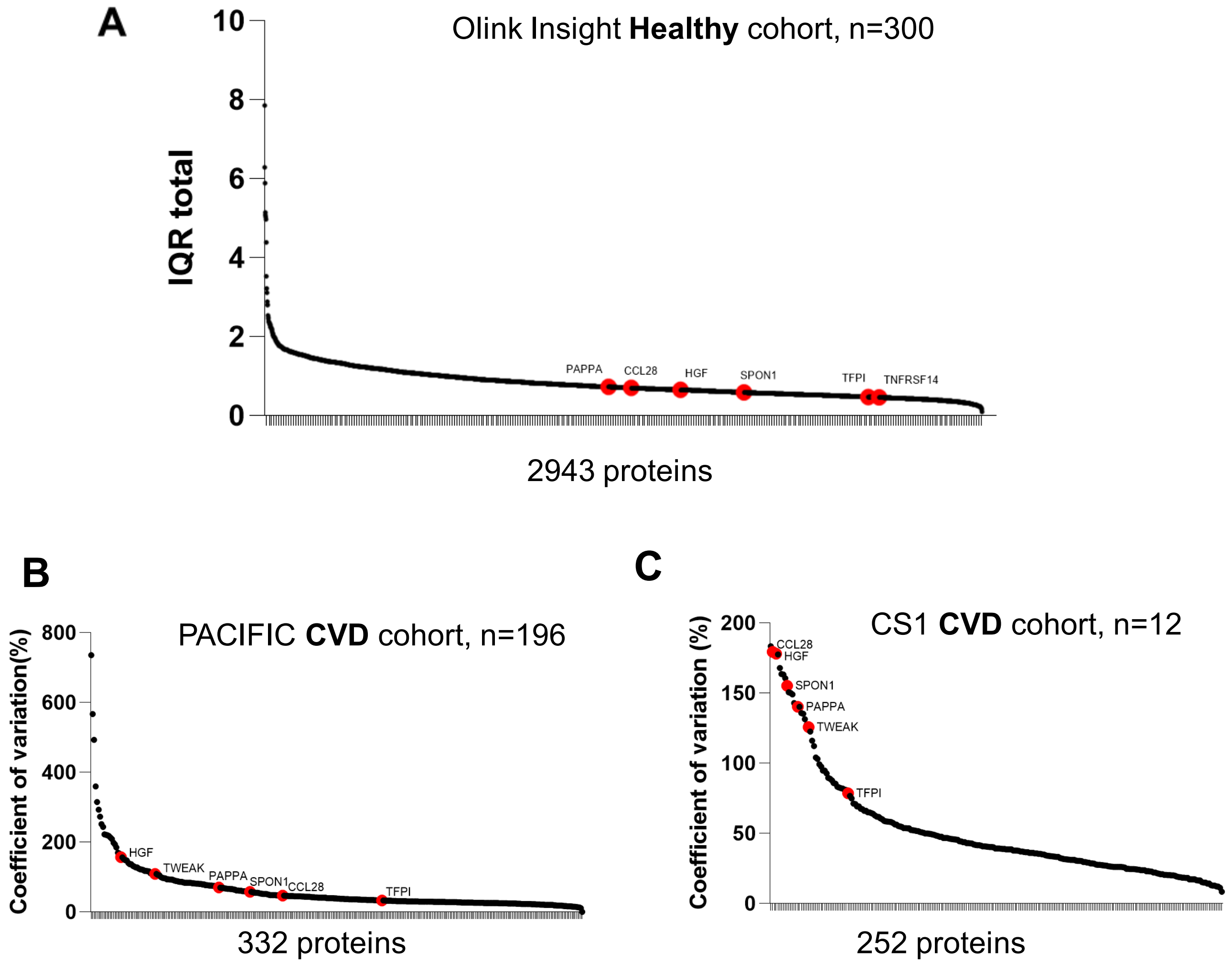


**Supplement Figure S2**. **Variability of proteins measured by IQR or coefficient of variation.**

A. Centred interquartile range (IQR) values downloaded from https://insight.olink.com/data-stories/normal-ranges for 2943 proteins. IQR values for biosignature proteins are highlighted in red are relatively low, i.e. this healthy cohort is unlikely to have a minority of participants with very different/high levels compared with the other participants.

B. Coefficient of variation in protein levels in the PACIFIC cohort where biosignature proteins are among the more variable proteins.

C. Coefficient of variation in protein levels in the CS1 cohort where biosignature proteins are among the more variable proteins.

### Supplementary Figures

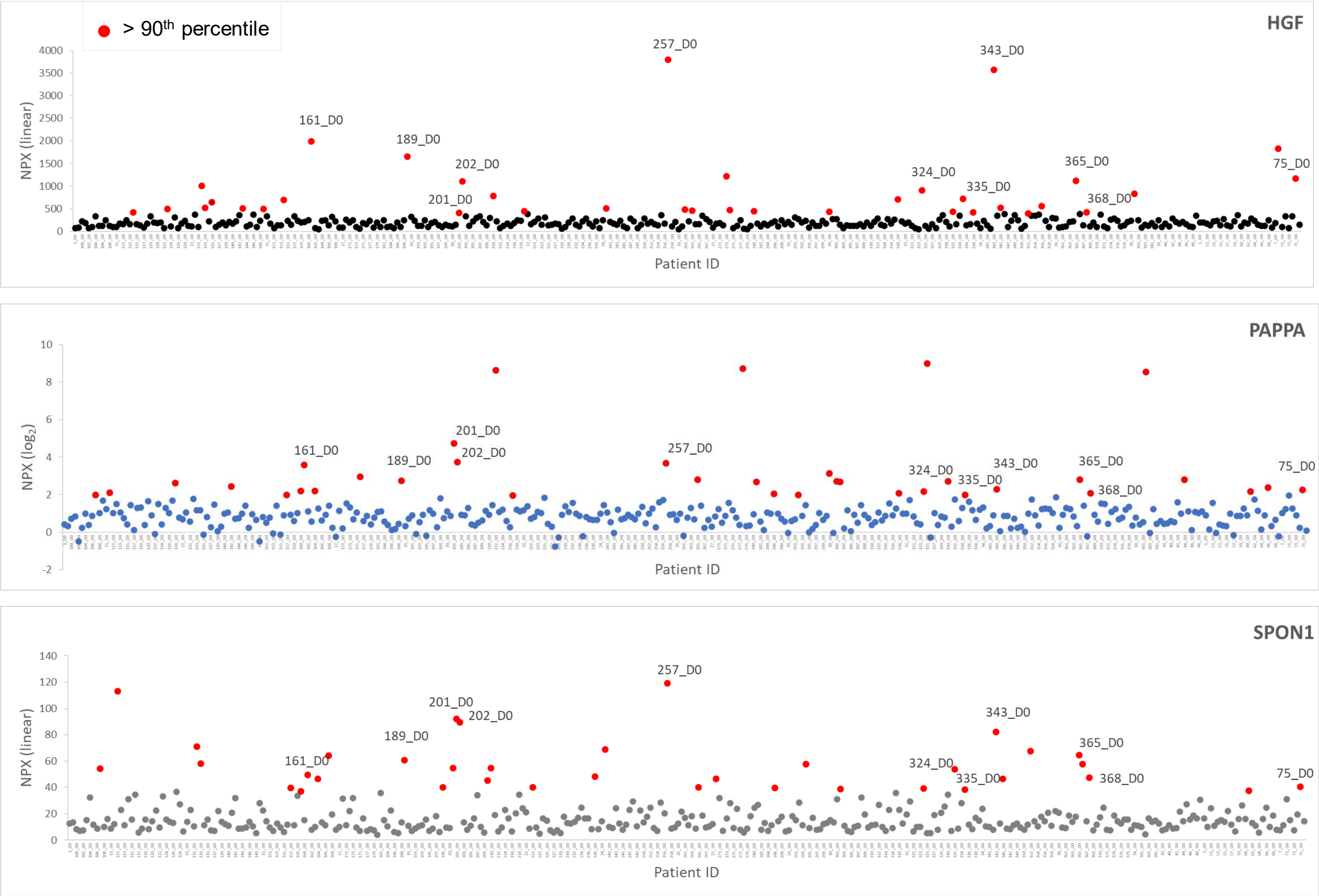


**Supplement Figure S3 . Identification of participants with the biosignature in the MGH COVID-19 study**
Data was downloaded from Filbin et al[27] and levels of HGF, PAPPA and SPON1 in plasma from participants at day 0 were used to identify participants with the biosignature. Participants with levels in the 90th percentile are displayed in red, while those below the 90th percentile are displayed in black (HGF), blue (PAPPA) and grey (SPON1). The 11 participants with plasma levels in the 90th percentile for HGF, PAPPA and also SPON1 are indicated by their ID number on the graph. Note that the MGH Boston COVID-19 NPX levels are 3 times higher than those in their reported dataset, this is taking into account the statement in the methods section that blood samples were diluted by a factor of 3 prior to analysis.

### Supplementary Figures

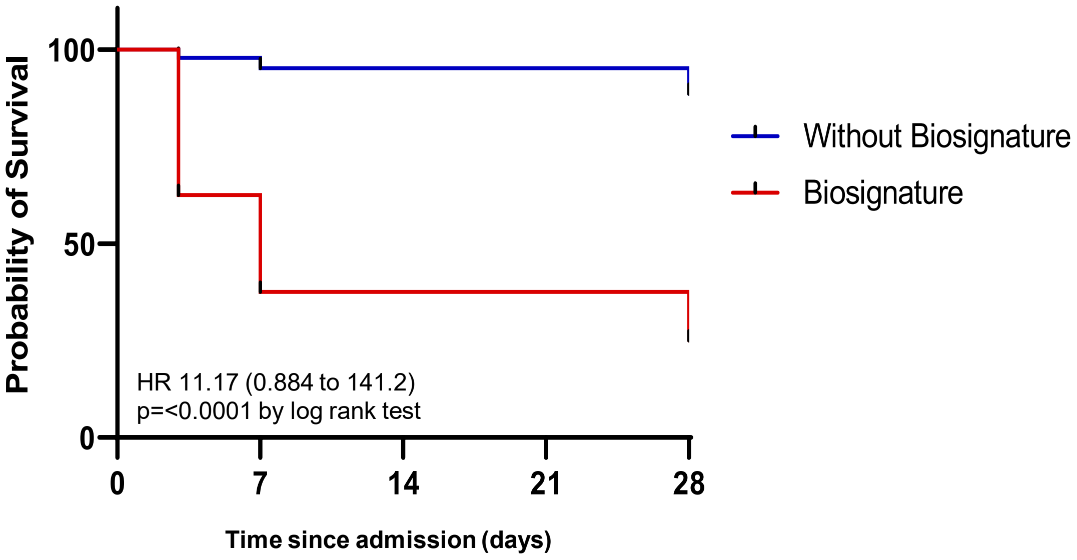


**Supplement Figure S4. Survival outcomes for patients with the biosignature in the MGH (COVID-19) cohort**
Kaplan-Meier curve displaying probability of survival (%) in MGH patients with the biosignature (11 patients) or without the biosignature (remaining 373 individuals). Shown are hazard ratios (HR, 95% CI and p value calculated by log rank test. Note that confidence intervals are large, due to the unbalanced groups and relatively small number of individuals in the biosignature group.

### Supplementary Figures

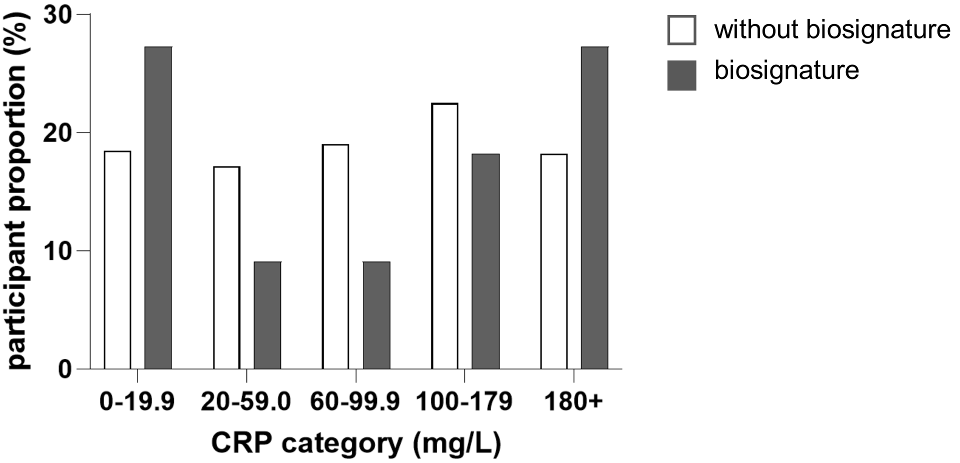


**Supplement Figure S5. CRP ranges in the MGH (COVID-19) cohort**
Proportions of participants in the different CRP categories, with or without the biosignature are displayed. Note that ranges of CRP levels were similar across the ranges, particularly at the lowest and highest ranges.

### Supplementary Figures

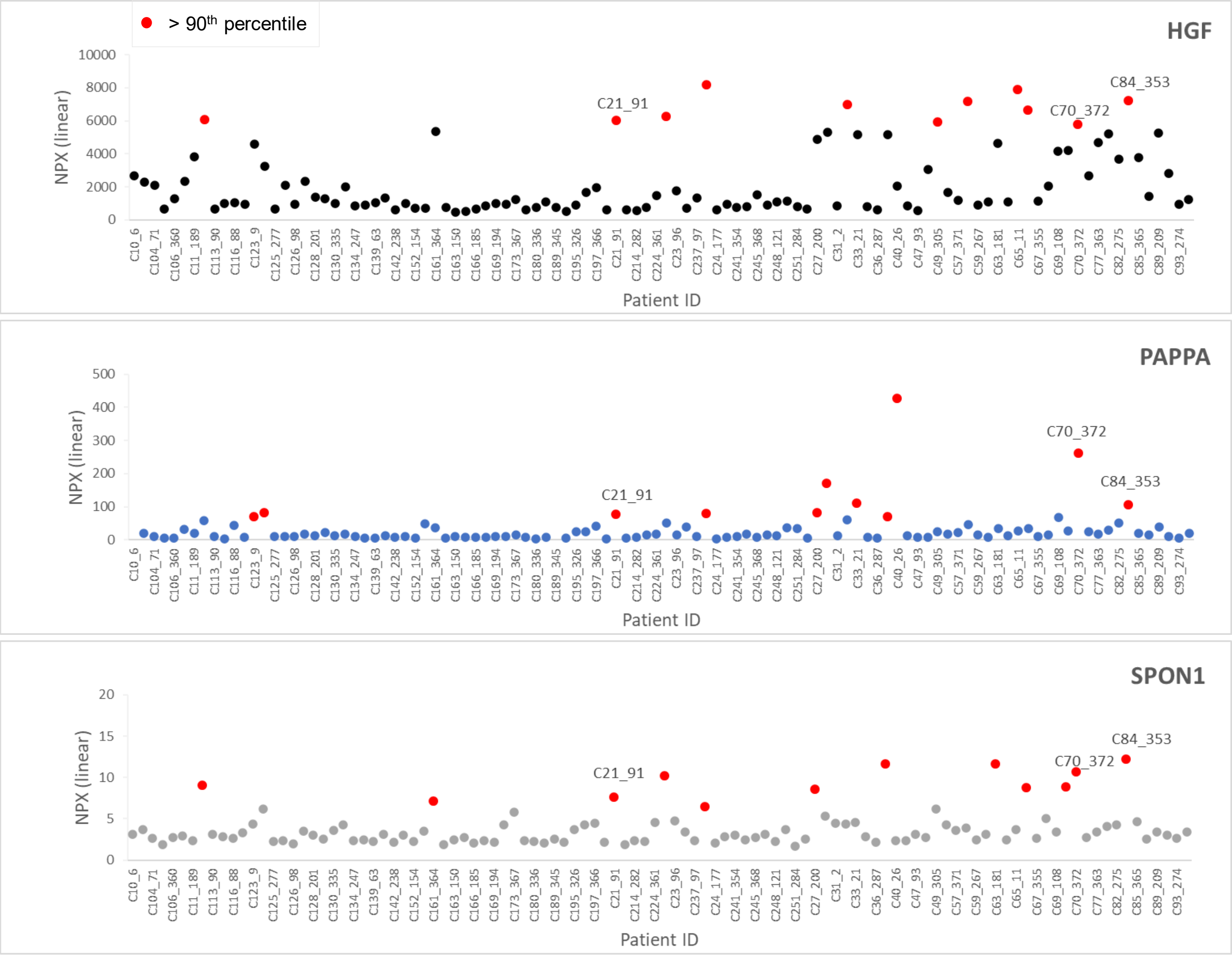


**Supplement Figure S6. Identification of participants with the biosignature in the Imperial College ESKD COVID-19 study**

Data was downloaded from Gisby et al[28] and levels of HGF, PAPPA and SPON1 in plasma were used to identify participants with the biosignature in subcohort A (first sample collected) are shown. Participants with levels in the 90th percentile are displayed in red, while those below the 90th percentile are displayed in black (HGF), blue (PAPPA) and grey (SPON1). The 3 participants with plasma levels in the 90th percentile for HGF, PAPPA and also SPON1 are indicated by their ID number on the graph. 2 of the participants with the biosignature tested positive for COVID-19 (C21 and C84) and 1 tested negative (C70).

### Supplementary Figures

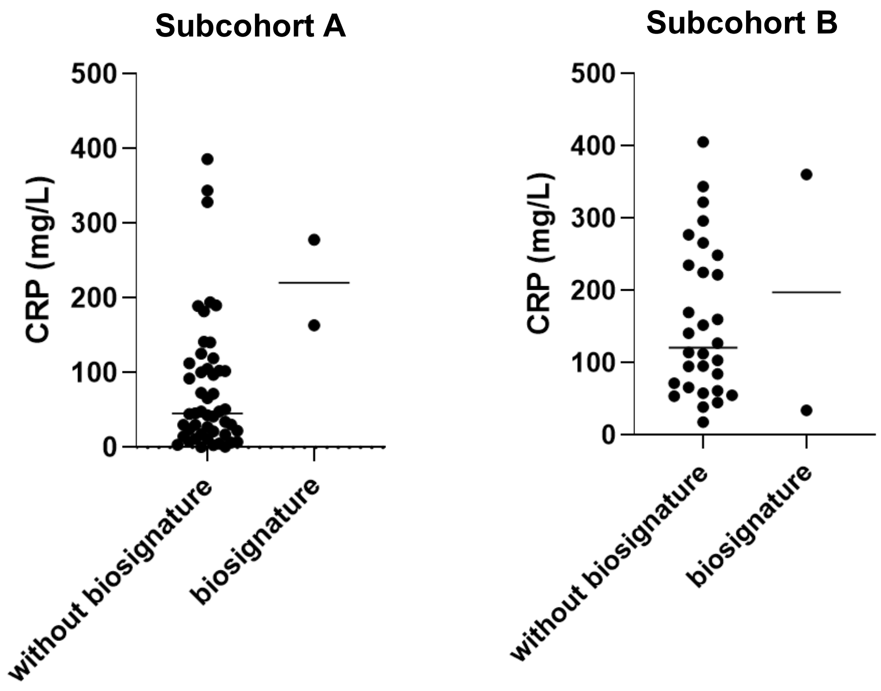


**Supplement Figure S7. CRP levels in the ESKD COVID-19 positive cohorts**
Levels of CRP in participants in subcohort A and subcohort B[28], with or without the biosignature are displayed. Note that participants with the biosignature (2 in each case) fall within the ranges of the whole cohort.

**Supplementary File S1 (Excel file attachment): List of proteins included in analysis and biosignature protein correlations with MPO, tryptase and syndecan-1**
Different cohorts and correlations between HGF, PAPPA, SPON1, CCL28, TFPI and TWEAK and mast cell (tryptase), neutrophil (MPO) and glycocalyx (syndecan-1) markers, where data is available on these protein levels. These are a series of tables showing the Spearman R and p values for each pairwise correlations.
